# Protective role of dietary antioxidant intake on long-term effects of extreme PM_2.5_ exposure on respiratory health

**DOI:** 10.1101/2024.12.11.24318880

**Authors:** Thara Govindaraju, Catherine L Smith, Jillian Ikin, Alice J Owen, Matthew Carroll, Brigitte M. Borg, Caroline X Gao, David Brown, David Poland, Shantelle Allgood, Michael J Abramson, Karen Walker-Bone, Tracy A McCaffrey, Tyler J Lane

**Affiliations:** School of Public Health and Preventive Medicine, Monash University, Melbourne, Victoria, Australia; Monash Rural Health Churchill, Monash University, Churchill, Victoria, Australia; Respiratory Medicine, Alfred Health, Melbourne, Victoria, Australia; Orygen, Centre for Youth Mental Health, The University of Melbourne, Parkville, Victoria, Australia; Department of Nutrition, Dietetics and Food, Monash University, Melbourne, Victoria, Australia

**Keywords:** landscape fires, respiratory health, PM_2.5_, dietary antioxidants, chronic cough, chronic phlegm

## Abstract

**Objective:** Evidence for the protective effects of dietary antioxidants on respiratory health following exposure to smoke from landscape fires is scarce. This study assessed whether the long-term respiratory effects of fine particulate matter (PM_2.5_) from the 2014 Hazelwood coal mine fire were moderated by dietary antioxidants, 8.5 years later.

**Methods:** We conducted a cross-sectional analysis of data collected 8.5 years after the mine fire from 282 residents of Morwell (a highly exposed town adjacent to the mine) and 166 residents from the nearby unexposed town of Sale. Coal mine fire-related PM_2.5_ was the primary exposure and meeting recommended dietary intake for antioxidants including vitamins (A, E and C), minerals (zinc, magnesium and selenium) and omega-3 fatty acids assessed through the Australian Eating Survey (AES) was the moderator. Respiratory symptoms were the primary outcome. Analyses used logistic regression models, adjusting for potential confounders.

**Results:** Vitamins A and E, magnesium, and zinc attenuated the association between fire-related PM_2.5_ and prevalence of chronic cough; three of the four (except magnesium) attenuated the association between fire-related PM_2.5_ and chronic phlegm. Omega-3 fatty acids attenuated the association between PM_2.5_ and COPD.

**Conclusion:** Meeting recommended intakes of vitamins, A and E, magnesium and zinc may provide protection against the long-term adverse effects of exposure to particulate matter from other medium duration but extreme exposure events. Advocating incorporation of fruits and vegetables in daily diet could offer more than the conventional health benefits due to their high antioxidant content.

**Highlights:**

➢ A coal mine fire exposed a regional Victorian community to extreme levels of PM_2.5_.
➢ We examined if dietary antioxidants mitigated PM_2.5_ effects on respiratory health.
➢ Vitamins A and E and Zinc reduced PM_2.5_ effects on chronic cough and chronic phlegm.

## 1. Introduction

On February 9, 2014, bushfires ignited the open cut brown coal mine adjacent to the Hazelwood power station in the Latrobe Valley in regional Victoria, Australia. The surrounding areas, especially the nearby town of Morwell, were covered in plumes of smoke and ash over a six-week period. In response to community concerns around possible health effects, the Victorian Department of Health commissioned a long-term study, which became the Hazelwood Health Study (Government of Victoria, 2014; Ikin et al., 2021).

There is substantial evidence linking PM_2.5_ to respiratory morbidity and mortality (Xing et al., 2016) as well as lung function impairment and increased risk of cardiopulmonary diseases (Dominici et al., 2006). Previous Hazelwood Health Study findings suggest smoke exposure from the coal mine has harmed respiratory health in the short and medium term (Holt et al., 2021; Johnson et al., 2019; Lane et al., 2023; Prasad et al., 2022), though there is some evidence of lung function recovery in the long term (Hemstock et al., 2024; Holt et al., 2024). Yet aside from the Hazelwood Health Study, there has been little evidence regarding the impact of PM_2.5_ from coal mine fires (Melody & Johnston, 2015). This is important given the unique nature of this mine fire, characterised by extreme but medium duration exposures to PM_2.5_ and related air pollutants (Franzi et al., 2011).

An optimal diet with higher fruit and vegetable intakes rich in antioxidants may reduce the harmful effects of air pollutants (Petriello et al., 2014; Whyand et al., 2018), including mitigating adverse effects of PM_2.5_ on respiratory health (Whyand et al., 2018). Elevated intracellular levels of reactive oxygen species (ROS) known as oxidative stress is implicated in the development of various chronic conditions including respiratory diseases. Under normal physiological conditions, antioxidants generated by endogenous mechanisms or ingested through diet can regulate concentrations of ROS (Pisoschi & Pop, 2015). In conditions such as asthma and chronic obstructive pulmonary disease (COPD), antioxidants reduce the oxidative stress that triggers respiratory inflammation (Andre et al., 2010).

Mechanistic pathways vary depending on the type of antioxidant and can include sequestration of free radicals, inhibition of free radical-producing enzymes, activation of endogenous antioxidant enzymes and prevention of lipid peroxidation and DNA damage (Carocho et al., 2018). Our previous work suggested that diet quality, and fruit and vegetable intake in particular, mitigated the effects of fire-related PM_2.5_ on respiratory symptoms (Govindaraju et al., 2024). This aligned with our *a priori* hypothesis that antioxidants in fruits and vegetables protected against or reduced the harmful effects of fire-related PM_2.5._

In this analysis, we explore this hypothesis further by examining intake of specific nutrients with antioxidant functions including minerals, vitamins and omega 3 fatty acids, commonly referred to as “dietary antioxidants”.

Key antioxidants, including vitamins A, C, and E, and minerals such as zinc, selenium and magnesium, are known to mitigate the oxidative damage implicated in conditions such as asthma, COPD, potentially reducing their severity and positively impacting lung function (Hoffmann & Berry, 2008; Pearson et al., 2005). Vitamin C is known to work both directly by donating electrons to free radicals to neutralise them, and indirectly by reactivating other free-radical scavengers such as alpha-tocopherol (vitamin E) and inhibiting free-radical-producing enzymes (Padayatty et al., 2003). Vitamin E, A and selenium, as components of selenoproteins and enzymes, act as peroxyl radical scavengers and maintain the integrity and hence the bioactivity of the long-chain polyunsaturated fatty acids in cell membranes (Traber & Atkinson, 2007). Zinc contributes to the synthesis of antioxidant enzymes, which neutralises free radicals and reduces inflammatory cytokines (Olechnowicz et al., 2018). Magnesium, a nutritionally significant mineral known for its role as a co-factor in more than 300 enzyme systems, regulates inflammation and oxidative stress (Ashique et al., 2023; Jahnen-Dechent & Ketteler, 2012). Apart from these micronutrients, omega-3 fatty acids, which are considered ‘essential’ since they cannot be made by humans and therefore must be part of the diet, are known immune modulators, especially of respiratory diseases relating to chronic inflammation (Knapp, 1995; Nursyifa Fadiyah et al., 2022).

Considering the overwhelming evidence on the role of antioxidants in reducing respiratory symptoms, coupled with a lack of evidence for their capacity to moderate the impacts of PM_2.5_, we investigated whether meeting recommended intakes of dietary antioxidants moderated the long-term effects of discrete but extreme PM_2.5_ exposure on respiratory health.

## 2. Methods

### 2.1 Study design and population

The study is a cross-sectional analysis of the 2022 follow-up to the Hazelwood Health Study’s (HHS) 2016/2017 Adult Survey (Ikin et al., 2021), which have both been described in detail previously (Govindaraju et al., 2024; Lane et al., 2024). In brief, the baseline Adult Survey cohort comprised 4,056 adults who had resided in either Morwell or the nearby but unexposed town of Sale during the mine fire. The 2022 follow-up 9 years after the mine fire (Lane et al., 2023, 2024), comprised 612 baseline cohort members from 2458 invited who had an email address or mobile phone number, had not participated in the HHS’s parallel Psychological Impacts stream (Carroll et al., 2022), and were not known to be deceased. Of those, 448 adults provided macro and micro-nutrient data which were included in these analyses. Adult Survey data were captured via the online REDCap platform (Harris et al., 2009, 2019), while the follow-up was captured entirely via REDCap.

### 2.2 Exposure – fire-related PM_2.5_

To determine individual-level exposure to fire-related PM_2.5_, we combined modelled temporal-spatial air pollution data with self-reported time-location diaries. The Commonwealth Scientific and Industrial Research Organisation’s (CSIRO) Oceans and Atmosphere unit used a chemical transport model to model mine fire-related PM_2.5_ concentrations in the Latrobe Valley and surrounding areas, incorporating variables such as wind direction, speed and temperature and the amount of PM_2.5_ released per unit mass of burning coal (Luhar et al., 2020). Time-location diaries were included in the Adult Survey, where cohort members provided their whereabouts at 12-hour intervals throughout the mine fire period (Ikin et al., 2021).

### 2.3 Moderator – dietary antioxidant intake

The follow up survey included the Australian Eating Survey (AES), a 120-item food frequency questionnaire developed by the University of Newcastle, Australia, which collected information on macro and micro-nutrient intake over the previous 6 months (Collins et al., 2014). Dietary antioxidants were selected from those available in the AES (Collins et al., 2014, 2015), based on prior evidence indicating they were protective against oxidative stress, especially with respect to respiratory diseases (Pisoschi & Pop, 2015). Each antioxidant was dichotomised based on daily recommended intake using Australian Nutrition Reference Value (NRV) cut-offs (Nutrient Reference Values for Australia and New Zealand, 2006). The values are listed in Table 1.

**Table 1.** Dietary antioxidants considered for analysis and their respective recommended dietary intakes per day.

| Antioxidant | Australian Nutrient Reference |  |
| --- | --- | --- |
|  | Values /day |  |
|  | Male | Female |
| Vitamin A (retinol equivalents) | 900µg | 700µg |
| Vitamin C | 45mg | 45mg |
| Vitamin E | 10mg | 7mg |
| Magnesium | 420mg | 320mg |
| Selenium | 70µg | 60µg |
| Zinc | 14mg | 8mg |
| Total long chain omega 3 fatty acids | 160mg | 90mg |

### 2.4 Outcome – respiratory symptoms

Respiratory symptoms and conditions were derived from a modified version of the European Community Respiratory Health Survey (ECRHS) III Short Screening Questionnaire (ECRHS Research Group, 2018) Self-reported symptoms and conditions included current wheeze, chest tightness, nocturnal shortness of breath, resting shortness of breath, current nasal symptoms, chronic cough, chronic phlegm, asthma and COPD post mine fire.

### 2.5 Confounders

All demographic factors were collected in the Adult Survey and included age (transformed with natural spline and three degrees of freedom to account for non-linear effects), sex, education (secondary up to year 10, secondary 11-12, or certificate/diploma/tertiary degree), and presence of pre fire asthma and COPD. Index of Relative Socioeconomic Advantage and Disadvantage (IRSAD) scores were derived from the 2016 Census of Population and Housing (Australian Bureau of Statistics, 2016) and matched to the participant’s residential area. Smoking behaviour was assessed both as smoking status (current, former, never (<100 lifetime cigarettes)) and pack years (e.g., 1 pack year equalled 20 cigarettes per day for 1 year) (Centers for Disease Control and Prevention (CDC), 2007).

### 2.6 Data Analysis

We utilised descriptive statistics to summarize the characteristics and outcomes of the cohort in both the exposed (Morwell) and non-exposed (Sale) groups. Differences between individuals residing in each location were evaluated using Fisher’s Exact tests for categorical variables and Kruskal-Wallis tests for continuous variables, with statistical significance set at p<0.05.

Multiple logistic regression models were developed to examine whether 10 µg/m^3^ changes in fire-related PM_2.5_ and dietary antioxidant intake were associated with respiratory symptoms. Each antioxidant was then treated as a moderator of fire-related PM_2.5_ effects (including an interaction term) in separate models for each respiratory outcome.

To increase statistical power and account for data assumed to be missing at random, we created multiple imputed datasets, with the number of imputations set equal to the proportion of records with missing data (n=8 for 7.14% of missing data). We then pooled the results according to Rubin’s rules (Buuren & Groothuis-Oudshoorn, 2011). Analyses were conducted via R (R Core team, 2023) and SPSS (Ver 28, IBM Corp, Armonk, NY). All R codes are made available on the Bridges repository (Lane, 2024).

## 2. Results

**Table 2** describes the key cohort characteristics and compares them by study location. Eight years after the mine fire, all self-reported respiratory symptoms were still more prevalent in Morwell than Sale. Prevalence of self-reported asthma was comparable between the two groups, whereas the prevalence of self-reported COPD was slightly higher in Sale compared to Morwell. Prevalence of current smokers was higher, while socio-economic status and educational attainment were lower in Morwell.

**Table 2.**
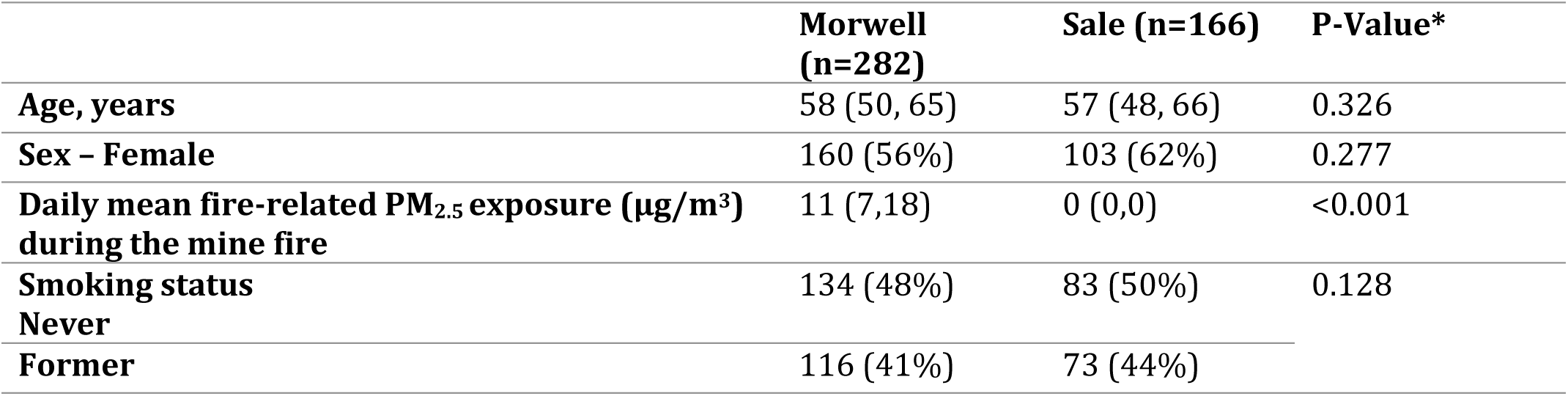
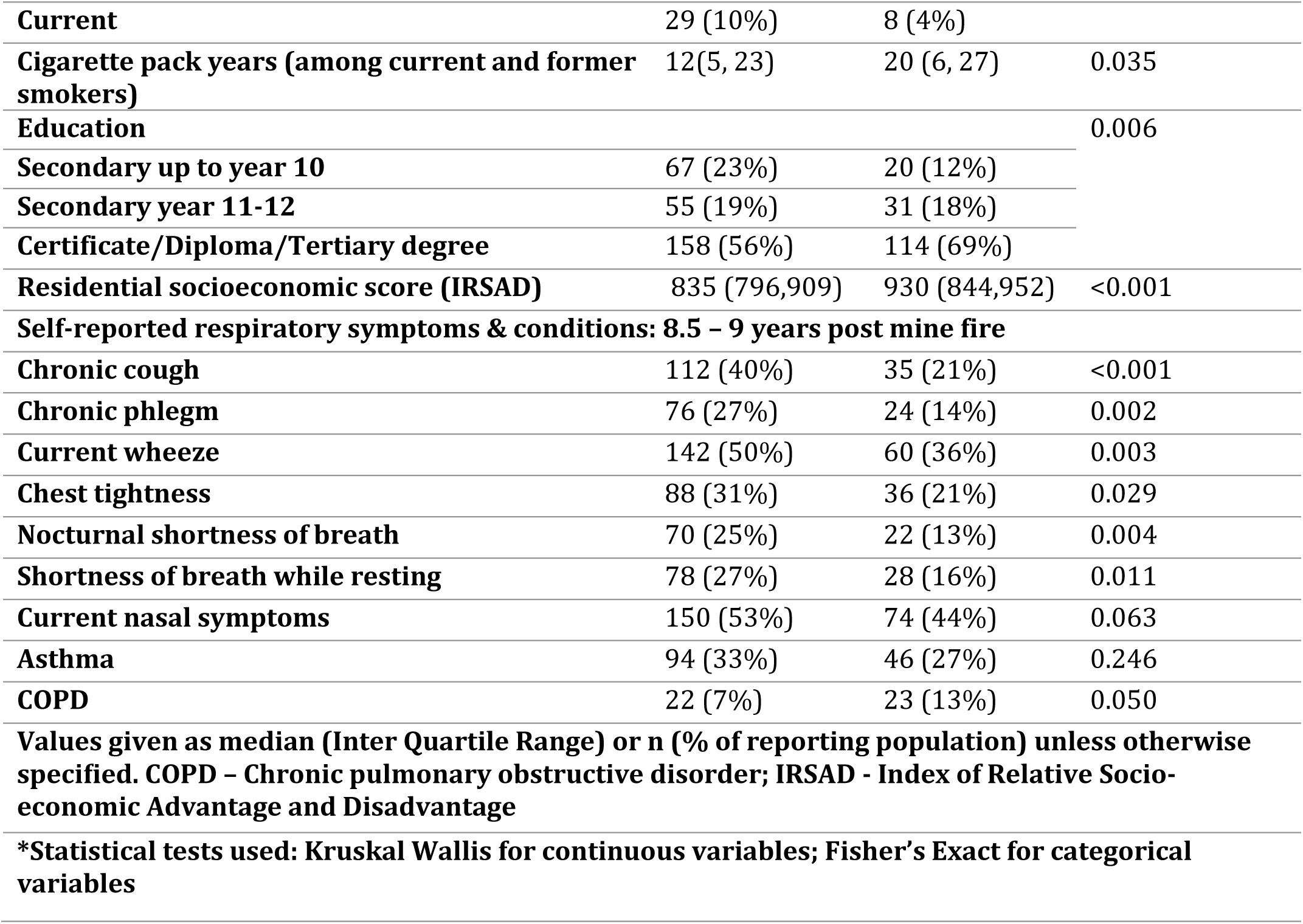
Participant characteristics across exposure (Morwell) and control (Sale) locations.

### Recommended intakes of dietary antioxidants among participants

Similar proportions of residents of Morwell and Sale met recommended daily intakes (RDI) of antioxidants based on Australian guidelines (Nutrient Reference Values for Australia and New Zealand, 2006), defined from here on as "adequate intake" (**Table 3).** The vast majority of participants met their recommended intakes for Vitamin C (>96%), as did large majorities for omega-3 fatty acids (>86%) and selenium (>75%).

**Table 3.**
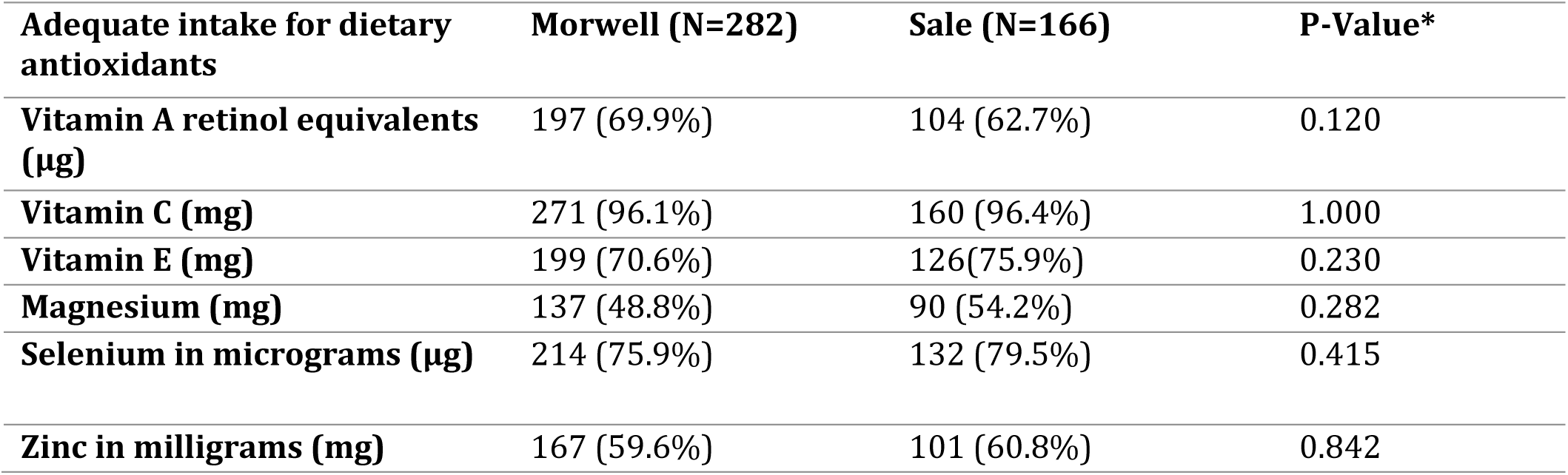
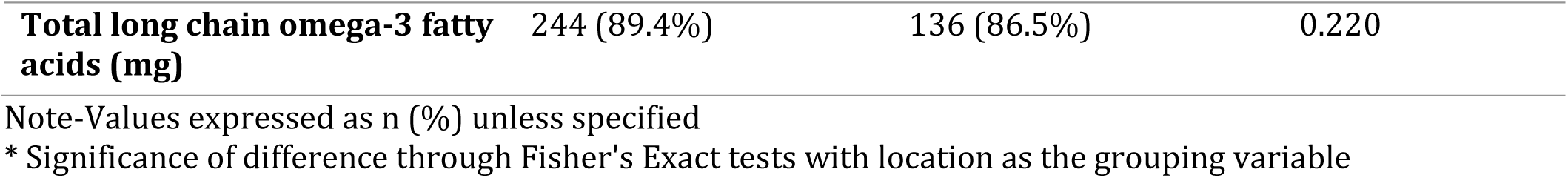
Percentage of participants meeting Australian guidelines for recommended daily dietary antioxidant intake by location.

| **Adequate intake for dietary antioxidants** | **Morwell (N=282)** | **Sale (N=166)** | **P-Value\*** |
| --- | --- | --- | --- |
| **Vitamin A retinol equivalents (µg)** | 197 (69.9%) | 104 (62.7%) | 0.120 |
| **Vitamin C (mg)** | 271 (96.1%) | 160 (96.4%) | 1.000 |
| **Vitamin E (mg)** | 199 (70.6%) | 126(75.9%) | 0.230 |
| **Magnesium (mg)** | 137 (48.8%) | 90 (54.2%) | 0.282 |
| **Selenium in micrograms (µg)** | 214 (75.9%) | 132 (79.5%) | 0.415 |
| **Zinc in milligrams (mg)** | 167 (59.6%) | 101 (60.8%) | 0.842 |
| **Total long chain omega-3 fatty acids (mg)** | **244 (89.4%)** | **136 (86.5%)** | **0.220** |

Note-Values expressed as n (%) unless specified
\* Significance of difference through Fisher's Exact tests with location as the grouping variable

### 3.2 Independent effects of dietary antioxidant intake and fire-related PM_2.5_ on respiratory health outcomes

Figure 1 summarises the association between fire-related PM_2.5_ and dietary antioxidant intake and respiratory symptoms in individual models, taking into account confounding factors. Fire-related PM_2.5_ exposure was associated with higher prevalence of self-reported chronic cough, current wheeze, and chronic phlegm. Among antioxidants, Vitamin A intake was associated with higher prevalence of asthma and Vitamin C was associated with higher prevalence of current nasal symptoms. Otherwise, there was no other association between antioxidants and respiratory health outcomes(Table S1).

**Fig. 1.**
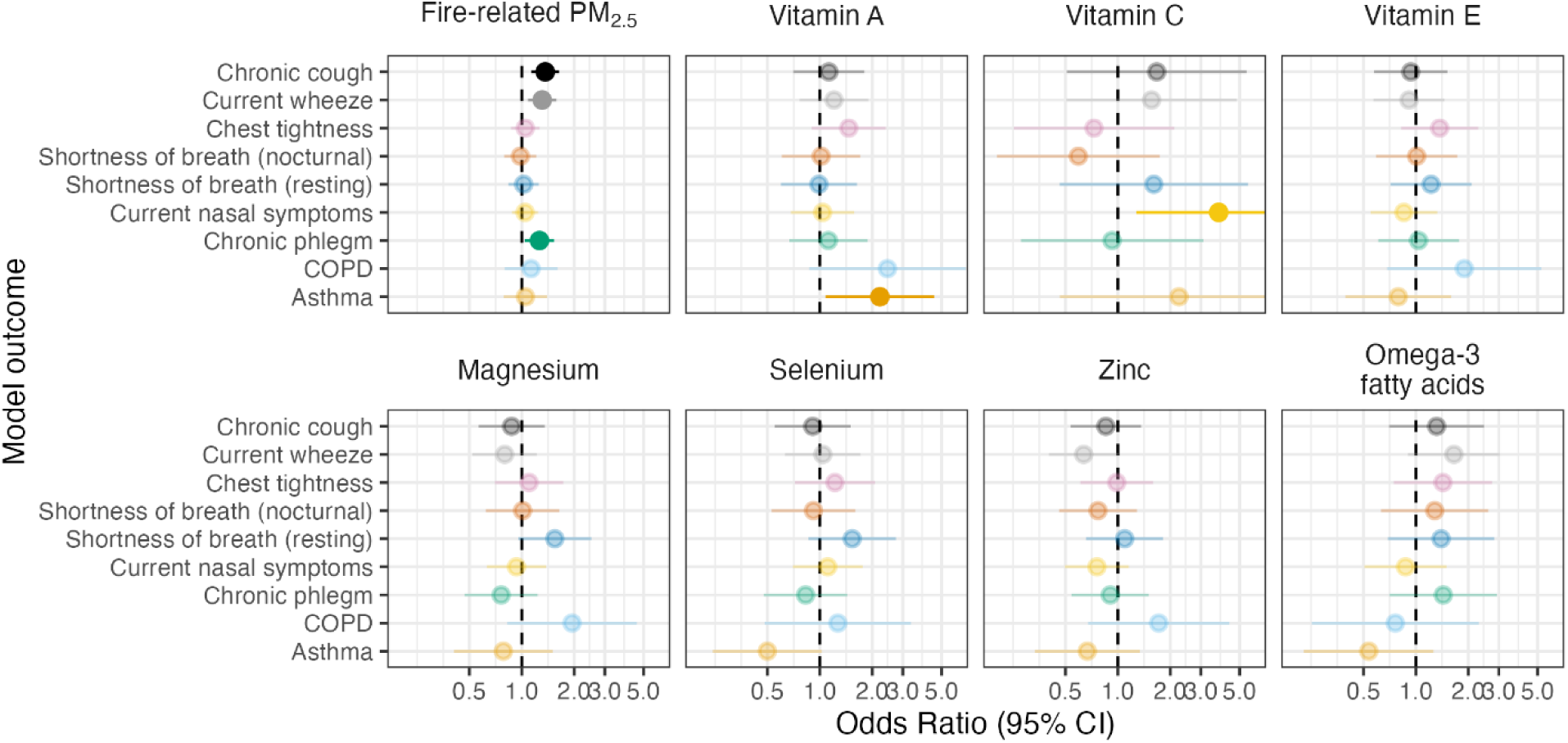
Forest plot showing the associations between 10 μg/m^3^ increases in fire-related PM_2.5_ and adequate intake of dietary antioxidants with the odds of reporting respiratory symptoms/conditions (*n* = 448). Odds Ratios were estimated from an imputed multivariable logistic regression controlling for age, sex, smoking, IRSAD, education and pre-fire asthma and COPD.

### 3.3 Interaction of dietary antioxidant intake and fire-related PM_2.5_ and respiratory health outcomes

Several antioxidants moderated the effects of PM_2.5_ on prevalence of chronic cough and chronic phlegm, including vitamins A and E, and zinc. Omega-3 fatty acids attenuated adverse effects of PM_2.5_ on COPD; this was the only moderated association for COPD and omega -3. These results are summarised in Figure 2 and Table S2.

**Fig. 2.**
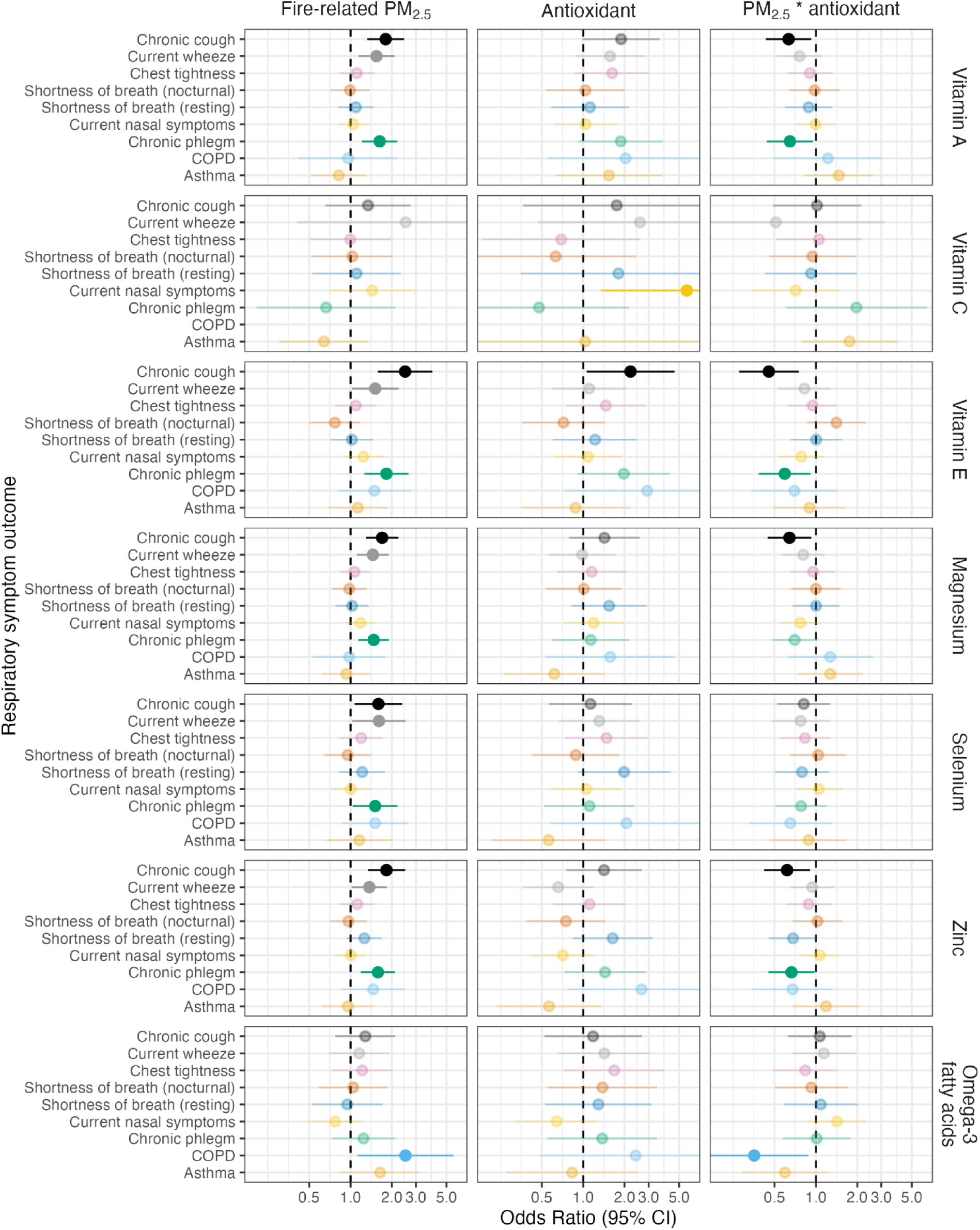
Forest plot showing the interactions between 10 μg/m^3^ increases in mine fire-related PM_2.5_ exposure and respiratory health outcomes, moderated by dietary antioxidant intake (*n* = 448). Each row of ORs were estimated from an imputed multivariable logistic regression fitting mine-fire PM_2.5_ exposure, dietary antioxidant intake, and interaction between PM_2.5_ exposure and dietary antioxidant intake controlling for age, sex, smoking, education, IRSAD score and pre-fire asthma and COPD. Note –The coefficients of vitamin C effects on COPD were not included due to the large confidence intervals, as detailed in Table S2. This was due to very low numbers of participants who had COPD and did not meet the RDI for vitamin C in Morwell (*n* = 1) and Sale (*n* = 0).

## 3. Discussion

When considered alongside our earlier study on the protective effects of foods high in antioxidants (Govindaraju et al., 2024), these new findings provide further evidence that dietary antioxidants may offer some protection against the respiratory harms of smoke from coal mine fires, particularly in relation to chronic cough, chronic phlegm and COPD. This new study is unique in its exploration of dietary antioxidants more directly.

Other recent studies have suggested a role for vitamins A, C and E in alleviating the harmful effects of air pollution on respiratory health outcomes, especially for chronic cough and phlegm (Whyand et al., 2018). While we found comparable results for vitamins A and E, we did not find comparable results with vitamin C. This is likely explained by the high proportion of participants (96%) meeting the daily recommended intake, leaving only *n* = 17 who consumed inadequate amounts of vitamin C, severely limiting statistical power to detect an interaction effect with this dichotomous outcome. Accordingly, the confidence intervals for vitamin C were the largest of any antioxidant group, regardless of whether the model included an interaction term.

Chronic cough merits some attention in terms of its effects on health and potential mechanisms to explain how it may be induced by fire-related PM_2.5_ and mitigated by dietary intake. The health effects of chronic cough include an association with worsened physical, mental, and social health due to sequalae such as exhaustion, restrictions on social engagement and isolation, family stress, and incontinence (Won & Song, 2021).It has persistently emerged as a respiratory symptom that is associated with exposure to PM_2.5_ from the Hazelwood coal mine fire (Govindaraju et al., 2024; Holt et al., 2021; Johnson et al., 2020; Lane et al., 2023; Prasad et al., 2022). However, treatment options are limited (Rouadi et al., 2022). Potential mechanisms underlying chronic cough may be illustrative of how antioxidants could be acting. PM_2.5_ and other air pollutants from chronic exposures to ambient sources, as well as discrete exposures to extreme events like 9/11 or earthquake rescues, may lead to cough hypersensitivity, or a lowered threshold to respond to stimuli with a cough (Jo & Song, 2019). The precise mechanism is not entirely clear, but these pollutants may cause neuropathology that upregulates neuroreceptors that can lower the threshold for a cough response (Jo & Song, 2019; Lv et al., 2016). Dietary antioxidants may modulate these receptors, providing relief from chronic cough (Guo et al., 2023) by reducing oxidative stress and inflammation, and may also improve airway inflammation to reduce hypersensitivity (Pisoschi & Pop, 2015). Apart from the antioxidant-vitamins examined in this study, flavonoids found in fruits and vegetables have antioxidant potential, and may modulate immune response, prevent inflammation, improve lung health and, by implication, alleviate related respiratory symptoms (Hassanpour & Doroudi, 2023). Additionally, Vitamins A, C and E are all indicated in cellular immunity and immune response mechanisms that reduce the frequency and severity of respiratory infections (Padayatty et al., 2003; Traber & Atkinson, 2007), which are common triggers for cough hypersensitivity.

Omega-3 fatty acids are thought to modulate respiratory diseases involving chronic inflammatory and infectious processes (Knapp, 1995). In this study, we found omega-3 intake mitigated PM_2.5_ effects on self-reported COPD in the post-fire period, yet we were unable to detect a main effect of fire-related PM_2.5_ on COPD, nor did previous Hazelwood Health Study analysis using self-reported data (Johnson et al., 2019). This raises the question: what effect is there to mitigate? It should be noted that these studies each relied on self-reported COPD which has been found to be unreliable. When measured more objectively through clinical tests, the Hazelwood Health Study found fire-related PM_2.5_ exposure was associated with spirometric COPD, i.e., spirometry consistent with COPD (Prasad et al., 2022). However, it is unclear why the current analysis was unable to detect a main effect between fire-related PM_2.5_ and COPD, yet an interactive effect. It is possible that this association is a chance finding. However, our earlier study (Govindaraju et al., 2024) observed an association between COPD and sauces and condiments, which was thought to be a statistical outlier and dismissed. While sauces and condiments are recognised causes of high sodium consumption and related chronic diseases burden among Australian adults (Bolton et al., 2020), they have also been postulated to contribute key nutrients (Whatnall et al., 2023) some of which are known to have antioxidant capacity *in vivo* (Whatnall et al., 2024; Andre et al., 2010). Omega-3 and lycopene and beta-carotene content found in fruits (tomato, in the case of sauces) and niacin found in Vegemite (Australian food spread made with brewer’s yeast) have found to exert synergistic effect and could possibly partly explain these findings (Townsend et al., 2023).

In contrast with other studies, we did not detect evidence of PM_2.5_ effect moderation by antioxidants for asthma or wheeze. While null results do not equate to no effect, it is worth expanding on other reasons why our results differ. Firstly, the antioxidant content that we analysed was gathered through a self-reported dietary intake tool, which is subject to recall bias and social desirability bias (Naska et al., 2017). More objective tests such as a serum antioxidant marker would provide more precise measurements and may lead to different results. Secondly, the antioxidant pathways are varied and complex. When a large number of these antioxidant nutrients are at work, the effects of one nutrient may negate the effect of the other. The unique characteristics of this community, which include residing in regional Australia and having overall poor diets (Govindaraju et al., 2024), may also affect how antioxidants interact with each other and both internal and external variables. On the other hand, it is also possible that several of these pathways require a synergistic effect. For instance, the role of zinc and selenium in DNA repair (Witkiewicz-Kucharczyk & Bal, 2006). Sub-optimal intake of one nutrient might affect the antioxidant potential of the other (Maggini et al., 2017; Townsend et al., 2023). The synergistic relationship between vitamin C and vitamin E is well documented, including in the regeneration of vitamin E to restore its antioxidant function and complementary functioning in lipid and aqueous cellular environments (Hamilton et al., 2000). This could mean that their combined effects enhance individual benefits and that insufficient levels of one could have an impact on the mechanistic pathway. Some clinical trials have explored the combined effects of vitamins C and E on health outcomes and found that co-supplementation results in improved markers of oxidative stress and inflammation by reducing inflammation and improving lung function (Hamilton et al., 2000), potentially relevant for conditions like chronic cough and asthma. Development of a validated cumulative antioxidant score to understand the effect of a combination of antioxidants as compared with a single dietary antioxidant could enhance the relevance of our findings. Lastly, we may have lacked statistical power to detect effects, largely because of the modest sample size, use of dichotomous moderator and outcome variables as well as interaction terms.

It is important to emphasise that the category of dietary antioxidants is not limited to the vitamins and minerals that have been explored in this study but also includes polyphenols, which have been studied extensively for their antioxidant potential(Hassanpour & Doroudi, 2023). Hence, there would be value in testing more holistic and comprehensive measures of antioxidants.

### 3.1 Strengths and limitations

This study has several strengths, including the utilization of a validated diet survey, modelled smoke/PM_2.5_ data offering individual-level exposure assessment, and the involvement of an established cohort. However, there are several limitations to consider. The analyses were cross-sectional and the relatively small sample size constrained statistical power. Consequently, significant associations arising from multiple comparisons should be interpreted cautiously. PM_2.5_ exposure relied on time-location diaries collected 2.5 years post-mine fire, which may have been inaccurate or even introduced recall bias and were unable to capture effects of some individual protective behaviours such as mask-wearing or staying indoors.

## Conclusions

Our findings suggest that recommended intakes of dietary antioxidants, especially vitamins A and E as well as magnesium and zinc, may protect against or alleviate the long-term consequences of smoke exposure from a coalmine fire, particularly chronic cough and chronic phlegm. This study contributes to the scarce body of evidence supporting the benefits of dietary antioxidants against the adverse effects of air pollution, although future studies would benefit from serum antioxidant-level assessments. Public health initiatives aimed at boosting fruit and vegetable intake could yield multiple benefits owing to their antioxidant content, especially given the escalating impacts of climate change on landscape fires and air quality.

## Supporting information

Supplement_Antioxidants and respiratory health.zip

## Data Availability

All data produced in the present study are available upon reasonable request to the authors

## Statements and Declarations

### Ethics

Monash University Human Research Ethics Committee approved this study as part of the Hazelwood Adult Survey & Health Record Linkage Study (Project ID: 25680; previously CF15/872 - 2015000389 and 6066)

### Availability of data and material

Hazelwood Health Study data are confidential and cannot be publicly shared. Analytical code has been archived on a public repository

## Acknowledgements

We thank the Latrobe Valley and wider Gippsland communities for their support and participation in the Hazelwood Health Study.

## Supplementary tables

**Table S1.** Regression models for dietary antioxidants and PM_2.5_ across respiratory health outcomes

**Table S2.** Regression models exploring the moderating role of dietary antioxidants between PM_2.5_ levels and respiratory health outcomes

