## Supplement_Antioxidants and respiratory health.zip for "Protective role of dietary antioxidant intake on long-term effects of extreme PM_2.5_ exposure on respiratory health": Supplement_Dietary antioxidants and respiratory health outcome_Preprint.pdf

#### Supplementary information

**Table S1.** Regression models for dietary antioxidants and $PM_{2.5}$ across respiratory health outcomes

| Respiratory health outcomes | Vitamin A | | Vitamin C | | Vitamin E | | Magnesium | | Selenium | | Zinc | | Omega 3 fatty acids | | PM2.5 | |
|---|---|---|---|---|---|---|---|---|---|---|---|---|---|---|---|---|
| | OR (95%CI) | p-value | OR (95%CI) | p-value | OR (95%CI) | p-value | OR (95%CI) | p-value | OR (95%CI) | p-value | OR (95%CI) | p-value | OR (95%CI) | p-value | OR (95%CI) | p-value |
| Chronic cough | 1.13 (0.70-1.80) | 0.618 | 1.67 (0.51-5.50) | 0.396 | 0.93 (0.58-1.51) | 0.784 | 0.87 (0.56-1.35) | 0.544 | 0.91 (0.55-1.51) | 0.784 | 0.85 (0.53-1.36) | 0.504 | 1.32 (0.70-2.46) | 0.391 | 1.36 (1.13-1.64) | 0.001 |
| Current wheeze | 1.21 (0.77-1.90) | 0.418 | 1.56 (0.51-4.82) | 0.436 | 0.91 (0.57-1.46) | 0.705 | 0.80 (0.52-1.22) | 0.296 | 1.04 (0.63-1.71) | 0.873 | 0.64 (0.40-1.01) | 0.053 | 1.65 (0.90-3.04) | 0.104 | 1.31 (1.08-1.58) | 0.005 |
| Chest tightness | 1.47 (0.90-2.39) | 0.125 | 0.73 (0.25-2.11) | 0.558 | 1.37 (0.82-2.28) | 0.225 | 1.10 (0.70-1.73) | 0.677 | 1.22 (0.72-2.07) | 0.454 | 0.99 (0.61-1.60) | 0.952 | 1.43 (0.75-2.74) | 0.281 | 1.05 (0.87-1.27) | 0.639 |
| Shortness of breath (nocturnal) | 1.02 (0.61-1.71) | 0.949 | 0.59 (0.20-1.74) | 0.341 | 1.01 (0.59-1.73) | 0.967 | 1.01 (0.62-1.64) | 0.965 | 0.92 (0.53-1.60) | 0.768 | 0.77 (0.46-1.29) | 0.318 | 1.28 (0.63-2.61) | 0.49 | 0.98 (0.79-1.21) | 0.86 |
| Shortness of breath (resting) | 0.99 (0.60-1.63) | 0.963 | 1.61 (0.46-5.59) | 0.453 | 1.22 (0.72-2.09) | 0.462 | 1.55 (0.96-2.50) | 0.076 | 1.53 (0.86-2.73) | 0.142 | 1.09 (0.66-1.82) | 0.728 | 1.40 (0.69-2.82) | 0.351 | 1.02 (0.84-1.25) | 0.816 |
| Current nasal symptoms | 1.13 (0.70-1.80) | 0.618 | 1.67 (0.51-5.50) | 0.396 | 0.93 (0.58-1.51) | 0.784 | 0.87 (0.56-1.35) | 0.544 | 0.91 (0.55-1.51) | 0.784 | 0.85 (0.53-1.36) | 0.504 | 1.32 (0.70-2.46) | 0.391 | 1.04 (0.88-1.24) | 0.624 |
| Chronic phlegm | 1.12 (0.67-1.88) | 0.669 | 0.93 (0.28-3.09) | 0.901 | 1.04 (0.61-1.77) | 0.896 | 0.76 (0.47-1.23) | 0.266 | 0.83 (0.48-1.44) | 0.522 | 0.90 (0.54-1.51) | 0.712 | 1.44 (0.70-2.93) | 0.318 | 1.26 (1.04-1.53) | 0.018 |
| Asthma | 2.21 (1.08-4.54) | 0.031 | 2.24 (0.46-10.84) | 0.314 | 0.79 (0.39-1.59) | 0.512 | 0.78 (0.41-1.51) | 0.463 | 0.50 (0.24-1.03) | 0.06 | 0.67 (0.33-1.34) | 0.253 | 0.54 (0.23-1.26) | 0.154 | 1.05 (0.79-1.39) | 0.742 |
| COPD | 2.44 (0.87-6.87) | 0.09 | 10506607.92 (0.00- Inf) | 0.988 | 1.90 (0.68-5.27) | 0.218 | 1.94 (0.83-4.57) | 0.128 | 1.27 (0.48-3.35) | 0.61 | 1.72 (0.67-4.37) | 0.256 | 0.76 (0.25-2.30) | 0.629 | 1.13 (0.80-1.60) | 0.494 |

Notes- OR-odds ratio /COPD- Chronic obstructive pulmonary disease; adjusted for age, sex, IRSAD and smoking /*significant at 0.05 levels

### Protective role of dietary antioxidant intake on long-term effects of extreme $PM_{2.5}$ exposure on respiratory health

**Table S2.** Regression models exploring the moderating role of dietary antioxidants between $PM_{2.5}$ levels and respiratory health outcomes; associations with $p<0.05$ **in bold**

#### Protective role of dietary antioxidant intake on long-term effects of extreme $PM_{2.5}$ exposure on respiratory health

| Dietary Antioxidants | Respiratory health outcomes | $PM_{2.5}$ | | Dietary antioxidants | | Interaction | |
|---|---|---|---|---|---|---|---|
| | | OR (95%CI) | P-Value | OR (95%CI) | P-Value | OR (95%CI) | P-Value |
| Vitamin A retinol equivalents (µg) | Chronic cough | 1.80 (1.32- 2.46) | **<0.001** | 1.89 (0.99- 3.60) | 0.054 | 0.63 (0.43- 0.93) | **0.019** |
| Vitamin C (mg) | | 1.34 (0.66- 2.74) | 0.419 | 1.75 (0.37- 8.38) | 0.48 | 1.02 (0.49- 2.14) | 0.959 |
| Vitamin E (mg) | | 2.50 (1.58- 3.95) | **<0.001** | 2.21 (1.06- 4.62) | **0.035** | 0.45 (0.28- 0.75) | **0.002** |
| Magnesium (mg) | | 1.70 (1.29- 2.23) | **<0.001** | 1.43 (0.79- 2.58) | 0.24 | 0.64 (0.44- 0.93) | **0.019** |
| Selenium in micrograms (µg) | | 1.60 (1.07- 2.38) | **0.021** | 1.13 (0.56- 2.27) | 0.731 | 0.82 (0.52- 1.27) | 0.367 |
| Zinc in milligrams (mg) | | 1.83 (1.34- 2.50) | **<0.001** | 1.42 (0.76- 2.66) | 0.275 | 0.62 (0.42- 0.91) | **0.014** |
| Total long chain omega 3 fatty acids (mg) | | 1.28 (0.78- 2.12) | 0.331 | 1.18 (0.52- 2.68) | 0.69 | 1.07 (0.63- 1.82) | 0.808 |
| Vitamin A retinol equivalents (µg) | Current wheeze | 1.55 (1.14- 2.10) | **0.005** | 1.57 (0.88- 2.83) | 0.129 | 0.76 (0.52- 1.11) | 0.154 |
| Vitamin C (mg) | | 2.52 (0.41-15.58) | 0.319 | 2.59 (0.47-14.31) | 0.274 | 0.51 (0.08- 3.18) | 0.47 |
| Vitamin E (mg) | | 1.52 (1.03- 2.24) | **0.037** | 1.11 (0.59- 2.08) | 0.754 | 0.83 (0.53- 1.28) | 0.388 |
| Magnesium (mg) | | 1.46 (1.11- 1.90) | **0.006** | 0.98 (0.57- 1.71) | 0.955 | 0.81 (0.56- 1.16) | 0.251 |
| Selenium in micrograms (µg) | | 1.61 (1.03- 2.52) | **0.036** | 1.31 (0.67- 2.58) | 0.433 | 0.77 (0.48- 1.26) | 0.298 |
| Zinc in milligrams (mg) | | 1.37 (1.02- 1.84) | **0.037** | 0.66 (0.37- 1.18) | 0.159 | 0.94 (0.65- 1.36) | 0.737 |
| Total long chain omega 3 fatty acids (mg) | | 1.16 (0.70- 1.91) | 0.571 | 1.43 (0.66- 3.08) | 0.366 | 1.14 (0.67- 1.96) | 0.622 |
| Vitamin A retinol equivalents (µg) | Chest tightness | 1.11 (0.83- 1.49) | 0.473 | 1.63 (0.87- 3.03) | 0.126 | 0.90 (0.62- 1.32) | 0.596 |
| Vitamin C (mg) | | 1.00 (0.50- 1.97) | 0.992 | 0.69 (0.18- 2.59) | 0.585 | 1.05 (0.52- 2.15) | 0.886 |
| Vitamin E (mg) | | 1.09 (0.78- 1.54) | 0.605 | 1.46 (0.75- 2.85) | 0.263 | 0.94 (0.63- 1.42) | 0.783 |
| Magnesium (mg) | | 1.07 (0.83- 1.38) | 0.603 | 1.15 (0.64- 2.07) | 0.628 | 0.96 (0.66- 1.39) | 0.808 |
| Selenium in micrograms (µg) | | 1.20 (0.83- 1.71) | 0.329 | 1.48 (0.74- 2.96) | 0.27 | 0.83 (0.55- 1.27) | 0.397 |
| Zinc in milligrams (mg) | | 1.12 (0.85- 1.48) | 0.429 | 1.11 (0.60- 2.05) | 0.73 | 0.89 (0.61- 1.29) | 0.525 |
| Total long chain omega 3 fatty acids (mg) | | 1.22 (0.73- 2.02) | 0.446 | 1.68 (0.73- 3.89) | 0.224 | 0.84 (0.49- 1.44) | 0.517 |

| | | | | | | | |
|---|---|---|---|---|---|---|---|
| Vitamin A retinol equivalents (µg) | Nocturnal shortness of breath | 0.99 (0.72- 1.38) | 0.967 | 1.04 (0.54- 2.02) | 0.91 | 0.98 (0.64- 1.49) | 0.921 |
| Vitamin C (mg) | | 1.03 (0.52- 2.05) | 0.93 | 0.63 (0.16- 2.43) | 0.502 | 0.94 (0.46- 1.94) | 0.875 |
| Vitamin E (mg) | | 0.77 (0.50- 1.18) | 0.229 | 0.72 (0.36- 1.44) | 0.353 | 1.41 (0.87- 2.30) | 0.166 |
| Magnesium (mg) | | 0.98 (0.74- 1.31) | 0.896 | 1.01 (0.54- 1.90) | 0.976 | 1.00 (0.66- 1.51) | 1 |
| Selenium in micrograms (µg) | | 0.95 (0.64- 1.41) | 0.814 | 0.88 (0.43- 1.83) | 0.736 | 1.04 (0.65- 1.66) | 0.873 |
| Zinc in milligrams (mg) | | 0.97 (0.71- 1.32) | 0.824 | 0.75 (0.39- 1.45) | 0.39 | 1.03 (0.68- 1.56) | 0.9 |
| Total long chain omega 3 fatty acids (mg) | | 1.05 (0.59- 1.86) | 0.877 | 1.38 (0.55- 3.46) | 0.488 | 0.93 (0.50- 1.71) | 0.805 |
| Vitamin A retinol equivalents (µg) | Shortness of breath while resting | 1.10 (0.81- 1.47) | 0.544 | 1.12 (0.58- 2.15) | 0.732 | 0.89 (0.60- 1.31) | 0.549 |
| Vitamin C (mg) | | 1.11 (0.53- 2.32) | 0.786 | 1.81 (0.35- 9.32) | 0.48 | 0.92 (0.43- 1.99) | 0.836 |
| Vitamin E (mg) | | 1.02 (0.71- 1.47) | 0.898 | 1.22 (0.60- 2.48) | 0.581 | 1.00 (0.65- 1.55) | 0.983 |
| Magnesium (mg) | | 1.03 (0.78- 1.36) | 0.849 | 1.55 (0.82- 2.90) | 0.176 | 1.00 (0.68- 1.49) | 0.988 |
| Selenium in micrograms (µg) | | 1.22 (0.83- 1.78) | 0.313 | 1.99 (0.91- 4.31) | 0.083 | 0.79 (0.51- 1.24) | 0.31 |
| Zinc in milligrams (mg) | | 1.26 (0.94- 1.69) | 0.118 | 1.64 (0.84- 3.20) | 0.147 | 0.68 (0.45- 1.02) | 0.062 |
| Total long chain omega 3 fatty acids (mg) | | 0.95 (0.52- 1.71) | 0.855 | 1.29 (0.53- 3.15) | 0.574 | 1.09 (0.58- 2.02) | 0.793 |
| Vitamin A retinol equivalents (µg) | Current nasal symptoms | 1.05 (0.80- 1.36) | 0.729 | 1.04 (0.62- 1.76) | 0.876 | 0.99 (0.71- 1.39) | 0.97 |
| Vitamin C (mg) | | 1.44 (0.70- 2.96) | 0.322 | **5.69 (1.34-24.07)** | **0.018** | 0.71 (0.34- 1.50) | 0.37 |
| Vitamin E (mg) | | 1.24 (0.89- 1.73) | 0.201 | 1.08 (0.61- 1.92) | 0.783 | 0.78 (0.53- 1.15) | 0.213 |
| Magnesium (mg) | | 1.18 (0.93- 1.51) | 0.172 | 1.19 (0.72- 1.97) | 0.488 | 0.77 (0.55- 1.08) | 0.127 |
| Selenium in micrograms (µg) | | 1.00 (0.72- 1.40) | 0.979 | 1.06 (0.58- 1.91) | 0.859 | 1.05 (0.72- 1.55) | 0.791 |
| Zinc in milligrams (mg) | | 1.01 (0.78- 1.30) | 0.969 | 0.71 (0.42- 1.21) | 0.208 | 1.07 (0.77- 1.49) | 0.696 |
| Total long chain omega 3 fatty acids (mg) | | 0.77 (0.49- 1.22) | 0.264 | 0.64 (0.32- 1.27) | 0.204 | 1.42 (0.87- 2.32) | 0.157 |
| Vitamin A retinol equivalents (µg) | Chronic phlegm | 1.63 (1.21- 2.20) | **0.001** | 1.88 (0.92- 3.81) | 0.082 | 0.65 (0.44- 0.95) | **0.028** |

| | | | | | | | |
|---|---|---|---|---|---|---|---|
| Vitamin C (mg) | | 0.66 (0.21- 2.14) | 0.492 | 0.48 (0.11- 2.16) | 0.337 | 1.97 (0.60- 6.45) | 0.261 |
| Vitamin E (mg) | | 1.83 (1.27- 2.64) | **0.001** | 1.98 (0.91- 4.27) | 0.083 | 0.59 (0.38- 0.92) | **0.018** |
| Magnesium (mg) | | 1.47 (1.14- 1.91) | **0.003** | 1.13 (0.60- 2.15) | 0.698 | 0.70 (0.48- 1.03) | 0.07 |
| Selenium in micrograms (µg) | | 1.51 (1.04- 2.20) | **0.031** | 1.11 (0.52- 2.37) | 0.78 | 0.78 (0.51- 1.21) | 0.264 |
| Zinc in milligrams (mg) | | 1.58 (1.19- 2.12) | **0.002** | 1.44 (0.73- 2.85) | 0.292 | 0.66 (0.45- 0.98) | **0.038** |
| Total long chain omega 3 fatty acids (mg) | | 1.25 (0.73- 2.12) | 0.42 | 1.38 (0.55- 3.45) | 0.496 | 1.01 (0.57- 1.78) | 0.966 |
| Vitamin A retinol equivalents (µg) | Asthma | 0.82 (0.52- 1.31) | 0.408 | 1.54 (0.63- 3.76) | 0.342 | 1.47 (0.83- 2.61) | 0.186 |
| Vitamin C (mg) | | 0.64 (0.30- 1.36) | 0.244 | 1.03 (0.16- 6.89) | 0.972 | 1.76 (0.78- 3.95) | 0.171 |
| Vitamin E (mg) | | 1.13 (0.68- 1.87) | 0.634 | 0.88 (0.35- 2.20) | 0.783 | 0.90 (0.49- 1.64) | 0.726 |
| Magnesium (mg) | | 0.93 (0.63- 1.39) | 0.734 | 0.62 (0.26- 1.44) | 0.263 | 1.27 (0.74- 2.20) | 0.387 |
| Selenium in micrograms (µg) | | 1.16 (0.67- 1.99) | 0.592 | 0.56 (0.22- 1.45) | 0.232 | 0.88 (0.47- 1.66) | 0.702 |
| Zinc in milligrams (mg) | | 0.96 (0.62- 1.48) | 0.84 | 0.56 (0.23- 1.36) | 0.202 | 1.18 (0.68- 2.07) | 0.552 |
| Total long chain omega 3 fatty acids (mg) | | 1.64 (0.85- 3.19) | 0.141 | 0.83 (0.28- 2.49) | 0.741 | 0.60 (0.29- 1.23) | 0.161 |
| Vitamin A retinol equivalents (µg) | COPD | 0.96 (0.42- 2.20) | 0.925 | 2.03 (0.55- 7.50) | 0.285 | 1.22 (0.49- 3.05) | 0.665 |
| Vitamin C (mg) | | 1.33 (0.00- Inf) | 1.000 | 11915717.68 (0.00- Inf) | 0.99 | 0.86 (0.00- Inf) | 1.000 |
| Vitamin E (mg) | | 1.49 (0.81- 2.77) | 0.202 | 2.92 (0.73-11.69) | 0.129 | 0.70 (0.34- 1.45) | 0.335 |
| Magnesium (mg) | | 0.98 (0.53- 1.80) | 0.946 | 1.57 (0.53- 4.64) | 0.409 | 1.27 (0.62- 2.61) | 0.516 |
| Selenium in micrograms (µg) | | 1.51 (0.87- 2.63) | 0.142 | 2.06 (0.58- 7.37) | 0.265 | 0.65 (0.33- 1.30) | 0.225 |
| Zinc in milligrams (mg) | | 1.46 (0.86- 2.50) | 0.161 | 2.66 (0.77- 9.20) | 0.121 | 0.68 (0.34- 1.33) | 0.257 |
| Total long chain omega 3 fatty acids (mg) | | 2.51 (1.12- 5.62) | **0.025** | 2.41 (0.48-12.04) | 0.283 | 0.36 (0.14- 0.88) | **0.026** |

Notes- OR-odds ratio /COPD- Chronic obstructive pulmonary disease; adjusted for age, sex, IRSAD and smoking /*significant at 0.05 levels
